# Exit strategies: optimising feasible surveillance for detection, elimination and ongoing prevention of COVID-19 community transmission

**DOI:** 10.1101/2020.04.19.20071217

**Authors:** Kamalini Lokuge, Emily Banks, Stephanie Davis, Leslee Roberts, Tatum Street, Declan O’Donovan, Grazia Caleo, Kathryn Glass

**Affiliations:** National Centre for Epidemiology and Population Health, The Australian National University, 62 Mills Road, Acton ACT 2601; Retired, National Centre for Epidemiology and Population Health, The Australian National University, 62 Mills Road, Acton ACT 2601

## Abstract

**Background:** Following successful implementation of strong containment measures by the community, Australia is now close to the point of eliminating detectable community transmission of COVID-19. We aimed to develop an efficient, rapid and scalable surveillance strategy for detecting all remaining COVID-19 community transmission through exhaustive identification of every active transmission chain. We also identified measures to enable early detection and effective management of any reintroduction of transmission once containment measures are lifted to ensure strong containment measures do not need to be reinstated.

**Methods:** We compared efficiency and sensitivity to detect community transmission chains through testing of: hospital cases; primary care fever and cough patients; or asymptomatic community members, using surveillance evaluation methods and mathematical modelling, varying testing capacities and prevalence of COVID-19 and non-COVID-19 fever and cough, and the reproduction number. System requirements for increasing testing to allow exhaustive identification of all transmission chains, and then enable complete follow-up of all cases and contacts within each chain, were assessed per million population.

**Findings:** Assuming 20% of cases are asymptomatic and all symptomatic COVID-19 cases present to primary care, with high transmission (R=2.2) there are a median of 13 unrecognised community cases (5 infectious) when a transmission chain is identified through hospital surveillance versus 3 unrecognised cases (1 infectious) through primary care surveillance. 3 unrecognised community upstream community cases themselves are estimated to generate a further 22-33 contacts requiring follow-up. The unrecognised community cases rise to 5 if only 50% of symptomatic cases present to primary care. Screening for asymptomatic disease in the community cannot exhaustively identify all transmission under any of the scenarios assessed. The additional capacity required to screen all fever and cough primary care patients would be approximately 2,000 tests/million population per week using 1/16 pooling of samples.

**Interpretation:** Screening all syndromic fever and cough primary care presentations, in combination with exhaustive and meticulous case and contact identification and management, enables appropriate early detection and elimination of community transmission of COVID-19. If testing capacity is limited, interventions such as pooling allow increased case detection, even given reduced test sensitivity. Wider identification and testing of all upstream contacts, (i.e. potential sources of infection for identified cases, and their related transmission chains) is critical, and to be done exhaustively requires more resources than downstream contact tracing. The most important factor in determining the performance of such a surveillance system is community participation in screening and follow up, and as such, appropriate community engagement, messaging and support to encourage presentation and compliance is essential. We provide operational guidance on implementing such a system.

**Funding:** No specific funding was received for this project, beyond the salary support the authors receive from their institutions and elsewhere. Professor Banks is supported by the National Health and Medical Research Council of Australia (Principal Research Fellowship 1136128).

## Background

Worldwide, countries are implementing measures to contain COVID-19. Internationally, empirical data demonstrates that all countries that have been able to exert substantive control of the COVID-19 pandemic have implemented strong containment and social distancing, combined with extensive surveillance.^1-3^ Australia, too, has achieved success with its strong containment measures. Community compliance with such measures, estimated at over 80%^4^, along with strong border controls and wider testing and management of identified cases and contacts, has meant that case numbers have dropped from a peak of over 450 cases, to less than 21 new cases per day throughout the country as of 17^th^ March 2020^5^.

Once strong containment, social distancing and other measures have been successful in reducing transmission, enhanced surveillance systems are needed which are capable of confirming that transmission has been eliminated from the community, and which can also detect and control early transmission resulting from reintroductions of disease into the community once containment measures begin to be lifted. Some modelling has raised concerns that where measures to control COVID-19 have been successful, there is likely to be a resurgence of community transmission when measures are lifted, and this cannot be contained by surveillance of hospital presentations^6^. Broader community-based surveillance and contact-tracing is a proven strategy for enabling early detection in infectious disease outbreaks^7^ and has the potential to address the resurgence demonstrated by modelling that only includes hospital–based case detection. Moreover, testing capacity, as well as case and contact follow up interventions, are continuing to improve internationally, meaning that more sensitive and sophisticated surveillance systems are increasingly possible.^8-10^

Currently, there is uncertainty on how best to achieve exhaustive identification of all transmission once prevalence is very low or undetectable through current surveillance strategies, and to detect and manage any reintroduction of community transmission once containment has been achieved. The World Health Organization, although recommending surveillance of influenza-like illness and severe respiratory illness, identifies these areas as a knowledge gap in the monitoring of community transmission.^2^ This lack of evidence hampers control efforts not only because it limits effective management of transmission, but because it contributes to a reluctance to implement strong containment and social distancing, due to fears that such measures would be required for extended periods.^11,12^ These fears can be addressed by a clear strategy for managing subsequent resurgences of community transmission once such measures are lifted, without the need for reinstitution of strong containment measures.

WHO representatives have stated that ongoing control of COVID-19 following lifting of containment and social distancing measures will require identification and elimination of all transmission chains.^13^ Exhaustive detection and elimination of community transmission chains is standard practice in management of high-risk pathogen outbreaks such as Ebola Virus Disease,^7,14^ and this experience provides useful guidance for the control of the COVID-19 pandemic.^1^ Although there are factors that contribute to COVID-19 being more difficult to control with such strategies (e.g. higher proportion of asymptomatic disease, shorter serial interval), there are also factors that mean it is more amenable to such measures than infections such as influenza (e.g. longer serial interval), which we have accepted as beyond our current capacity to eliminate.

This study evaluates surveillance options and proposes an efficient, feasible and scalable strategy that will:

a. provide corroborating evidence that community transmission has been eliminated through strong containment measures when notified cases are declining;
b. identify and manage any early resurgence in transmission once these measures are lifted;
c. address current constraints in surveillance such as limited testing capacity; and
d. identify other response priorities that are critical to enhanced surveillance.

A range of options are assessed, and efficient strategies are recommended, including a summary of the structure and requirements for such a public health surveillance system. Given constraints on testing, we provide a range of scenarios and related testing and follow-up requirements to allow the proposed strategy to be scaled up and tailored to available capacity. This includes assessing test performance for varying COVID-19 prevalence, and assessing the benefits and costs of pooling, a strategy now being implemented in some settings with limited test materials^15,16^. Although increasing the number that can be tested, pooling may decrease sensitivity^17^, and is of varied efficiency depending on disease prevalence, thus we sought to assess the impact of such considerations.

## Methods

Based on current and potentially effective surveillance strategies from our rapid review, we compare the following three candidate groups for surveillance:

- Group 1: Patients with pneumonia presenting to hospitals
- Group 2: Patients with fever plus cough presenting to primary care (either general practices or specialist fever clinics). Fever and cough is recommended by WHO as a general syndromic surveillance case definition^2^, and has been reported as commonly occurring symptoms in patients with COVID-19^18^. We therefore used this as the syndromic case definition for identification of suspected symptomatic disease in the community.
- Group 3: Asymptomatic community members who may have a higher level of unprotected exposure to the community e.g. supermarket and delivery workers, transport workers,^19^ essential service staff living in families with children who attend school, and teachers (if schools are open).

The following were estimated for these three groups:

- The estimated number of cases in the community within the same transmission chain as the detected case at the time the detected case presents.
- The sensitivity and efficiency of testing these groups for a range of testing capacities and varying prevalence of non-COVID-19 fever and cough patients in primary care, and for varying prevalence of COVID-19 in sentinel populations.
- The feasibility of surveillance both qualitatively and quantitatively, including the system requirements for increasing testing and follow-up in these groups for a population prevalence of COVID-19 of 1/20,000. This prevalence was calculated with the following assumptions: Iceland had the highest level of community-based screening globally as at 23/03/20, and a similar epidemiology to Australia. Their general population overall prevalence is 0.86% (48/5571). Prevalence in health care system testing was 425/4197=10%, with 34% of this travel related. We assumed 0.005% weekly period prevalence in community, which is 1/3 of that, since community transmission in Australia has decreased substantially since this time.

Gains from enhanced community surveillance are estimated for two scenarios:

1. High levels of transmission, in which surveillance is implemented in a context of limited or no containment measures and therefore the reproductive number is assumed to be 2.2,^20^ (i.e. the situation if most containment measures are lifted following successful elimination)
2. Low levels of transmission, in which surveillance is conducted following lifting of some containment measures that have reduced transmission, with the reproductive number assumed to consequently rise above one to 1.2.

Estimated cases in the community were calculated using a stochastic SEIR model, assuming a four-day incubation period before symptom onset, with transmission occurring during the final day of this period. We assumed three levels of severity: severe cases (20% of all symptomatic cases)^21^ who are hospitalised, asymptomatic cases (20% of all cases^22^, with the remaining 60% of cases experiencing mild or moderate disease and not requiring hospitalisation. In sensitivity analyses, we considered higher rates of asymptomatic infection, ranging up to 50%.^23^ As hospital surveillance is currently restricted to cases with pneumonia, we assumed a 7-day delay from symptom onset until presentation at hospital in severe cases,^24^ while a proportion of mild and moderate cases present to primary care two days after developing symptoms. We assume that cases are only infectious for three days (including one day of pre-symptomatic transmission) to replicate serial intervals of around 4-6 days.^24^

Sensitivity and efficiency of testing strategies was evaluated using the following assumptions to calculate precision of point prevalence estimates and case detection capacity per million population:

- Random sampling of presentations in each group.
- Online syndromic surveillance data in Australia (FluTracker) provides self-reported prevalence of fever and cough on a weekly basis. From 2019 data,^25^ we expect a peak population prevalence of cough and fever of 3%, with 2% across much of winter and 1% outside winter. With extensive social distancing measures, prevalence of fever and cough due to all causes may decrease by 40%,^26^ therefore we also assessed 0.6% non-COVID fever and cough prevalence.
- We compared primary care presentation rates of 50% and 100% of patients with fever and cough in the community.

We also assessed overall test performance (negative and positive predictive values and false negative and false positive rates) under a range of prevalence values and test performance in terms of sensitivity and specificty. Benefits of pooling of samples were considered by estimating the number of tests required at a given prevalence. We also assessed the relative benefits compared to costs of pooling if pooling were to result in a reduction in sensitivity by comparing a situation of high test sensitivity and no pooling (and therefore reduced number of patients tested) with a range of lower sensitivities and pooling (and therefore a higher number tested but also a higher number of false negatives).

## Results

### Gains from early detection

Figure 1 presents the number of infected people in the community within a single transmission chain at the time that there is one case detected by primary care (left) or hospital (right) surveillance, assuming hospital detections are severe cases with pneumonia and 50% of all symptomatic cases present to primary care with cough and fever. Primary care surveillance results in fewer undetected cases in the community to be identified through contact tracing: under the high transmission scenario (reproductive number of 2.2) there are a median of 13 infected people in the community when a severe case is detected at the hospital compared to a median of 5 infected people in the community when there is a detection through primary care. Under the low transmission scenario (reproductive number of 1.2) there are a median of 4 infected people in the community when a severe case is detected at the hospital compared to a median of 3 infected people in the community when there is a detection through primary care. If all symptomatic cases present to primary care, there are just 3 infected people to be traced under high transmission and 2 infected people under low transmission. Conversely if 50% of cases are asymptomatic and only 50% of symptomatic cases present to primary care, there are a median of 6 infected people in the community when there is a detection through primary care at high transmission, and a median of 3 infected people in the community at low transmission.

**Figure 1:**
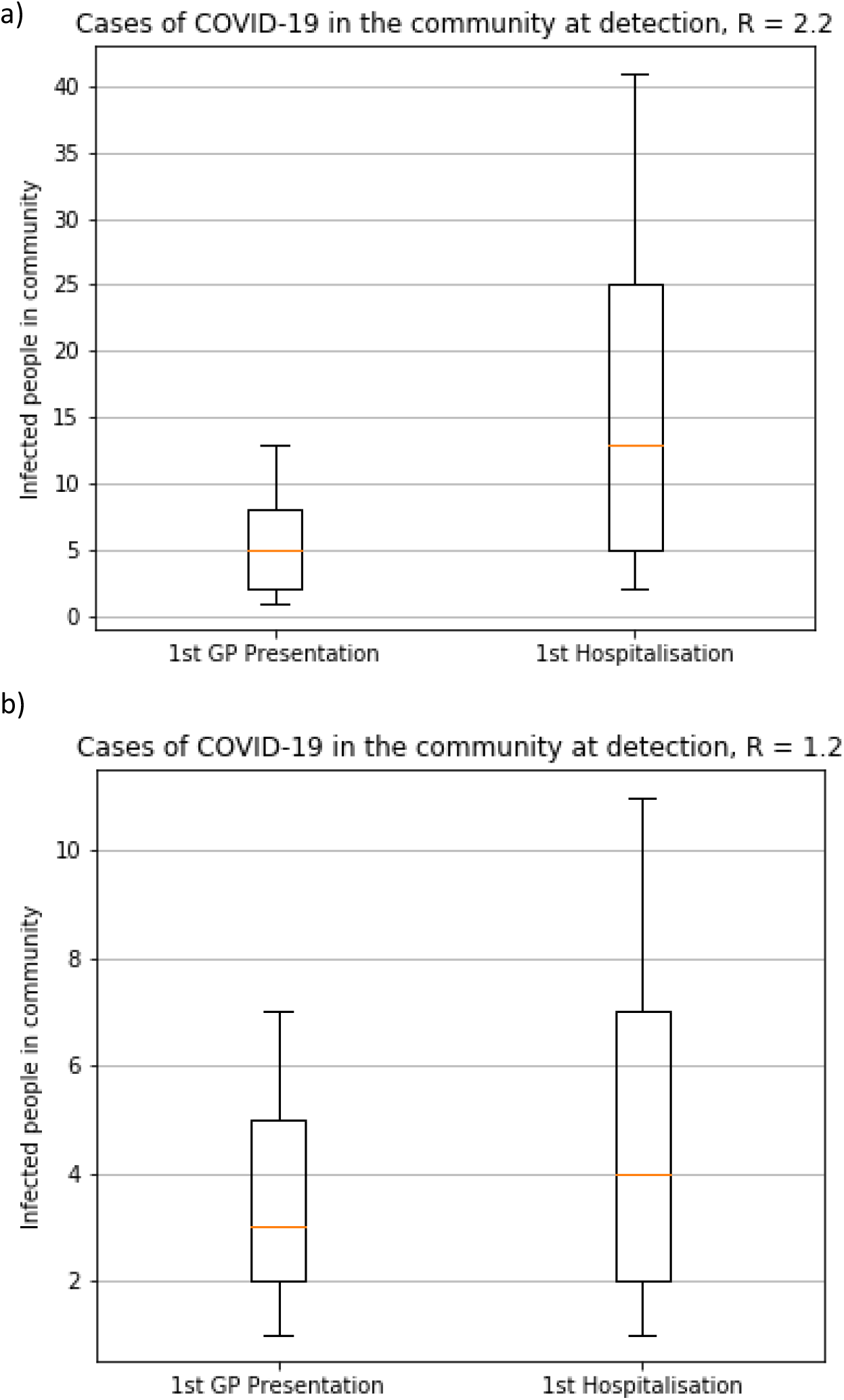
Cases of COVID-19 in the community at detection for R or 2.2. or 1.2 for community detection compared with hospital detection of cases. Number of infected people in the community when there is one detected case in general practice (left) or at hospital (right) for high and low reproductive numbers (R). For each scenario, the box shows the median, 25% and 75% of the distribution of number of cases, while the interval indicates the 10% and 90% limits.

### Efficiency of testing

#### (a) Fever and cough testing in primary care

Table 1 assesses, for a weekly incidence of 1/20,000 cases in the community,^1^ the estimated prevalence and number of community cases of COVID-19 missed by screening, given varied levels of non-COVID-19 fever and cough in the community, and varied levels of testing in these patients. Under the most likely assumptions outside of winter (1% fever and cough prevalence), exhaustive testing of all fever and cough patients would be possible with 10,000 samples per million population per week. If COVID-19 cases represent 0.1-1% of these fever and cough patients, population testing could be achieved with 800-2,200 tests per million population by pooling of samples in batches of 16 (Table 3).

**Table 1:**
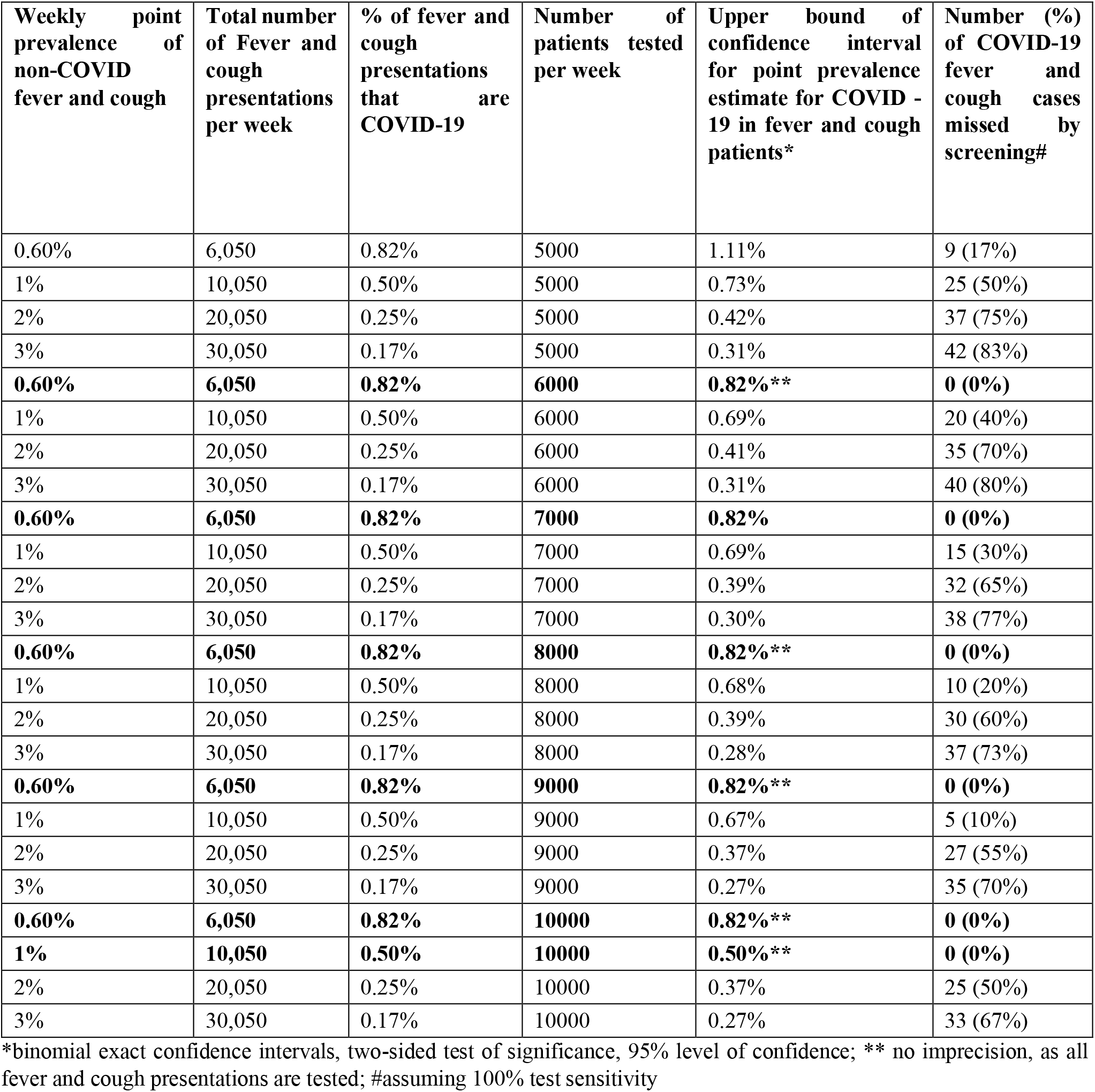
Number of COVID-19 community cases missed under varied non-COVID-19 fever and cough prevalence and testing levels per million population per week, assuming weekly period prevalence of COVID-19 related fever and cough in community is 1/20,000 (50/million), and all patients present for testing.

The above results are primarily influenced by the prevalence of non-COVID-19 fever and cough in the general community. Even at much higher or lower prevalence of COVID-19, the overall conclusions remain the same. The main impact of varied community prevalence is on precision (confidence interval size around prevalence estimate), on absolute numbers of cases missed and on efficiency gains through pooling of tests. However, when all fever and cough patients present and testing is exhaustive, the number of COVID-19 community cases missed by screening remains at 0 whatever the COVID-19 community prevalence.

#### (b) Testing of asymptomatic groups with high-risk contacts

Table 2 assesses the estimated prevalence and number of community cases of COVID-19 missed by screening, given varied testing and prevalence of COVID-19 infection in an asymptomatic sentinel population. For all scenarios assessed, the majority of asymptomatic cases are missed by random screening of sentinel populations

**Table 2:** Number and percentage of asymptomatic COBID-19 infections missed on screening, for a range of prevalence of asymptomatic COVID-19 infection and testing levels, per million population.

| **Prevalence of asymptomatic COVID-19 infection** | **Number of tests conducted** | **Upper bound of confidence interval for point prevalence estimate for COVID-19 in asymptomatic, high-risk group\*** | **Number of cases in screened population** | **Number of COVID-19 asymptomatic infections missed by screening** | **% of asymptomatic COVID-19 infections in populations that are missed during screening** |
| --- | --- | --- | --- | --- | --- |
| 0.79% | 10,000 | 0.98% | 7900 | 7821 | 99.0% |
| 0.48% | 10,000 | 0.64% | 4800 | 4752 | 99.0% |
| 0.24% | 10,000 | 0.36% | 2400 | 2376 | 99.0% |
| 0.16% | 10,000 | 0.26% | 1600 | 1584 | 99.0% |
| 0.10% | 10,000 | 0.18% | 1000 | 990 | 99.0% |
| 0.79% | 15,000 | 0.95% | 7900 | 7782 | 98.5% |
| 0.48% | 15,000 | 0.60% | 4800 | 4728 | 98.5% |
| 0.24% | 15,000 | 0.33% | 2400 | 2364 | 98.5% |
| 0.16% | 15,000 | 0.24% | 1600 | 1576 | 98.5% |
| 0.10% | 15,000 | 0.16% | 1000 | 985 | 98.5% |
| 0.79% | 20,000 | 0.92% | 7900 | 7742 | 98.0% |
| 0.48% | 20,000 | 0.59% | 4800 | 4704 | 98.0% |
| 0.24% | 20,000 | 0.32% | 2400 | 2352 | 98.0% |
| 0.16% | 20,000 | 0.23% | 1600 | 1568 | 98.0% |
| 0.10% | 20,000 | 0.15% | 1000 | 980 | 98.0% |
| 0.79% | 25,000 | 0.91% | 7900 | 7703 | 97.5% |
| 0.48% | 25,000 | 0.57% | 4800 | 4680 | 97.5% |
| 0.24% | 25,000 | 0.31% | 2400 | 2340 | 97.5% |
| 0.16% | 25,000 | 0.22% | 1600 | 1560 | 97.5% |
| 0.10% | 25,000 | 0.15% | 1000 | 975 | 97.5% |

**Table 3:** Proportion of COVID-19 infections missed per 10,000 population, with and without pooling, and accounting for varied sensitivities with pooling (given test specificity of 99.9% throughout and sensitivity of 99.9% in non-pooled tests)

| row | prevalence | Exhaustive testing of 10,000 individuals using 1/16 pooling |  |  |  |  | Individual testing of the same number of people as in column 3 |  |
| --- | --- | --- | --- | --- | --- | --- | --- | --- |
|  |  | Total tests | sensitivity | NPV | Number with disease | Percentage with disease missed | Percentage with diseased missed (sensitivity in column 4) | Percentage with disease missed (sensitivity of 99.9%) |
| 1 | 0.05% | 715 | 99.90% | 100.00% | 5 | 1% | 93% | 93% |
| 2 | 0.05% | 711 | 95.00% | 100.00% | 5 | 5% | 93% | 93% |
| 3 | 0.05% | 707 | 90.00% | 99.99% | 5 | 10% | 94% | 93% |
| 4 | 0.05% | 699 | 80.00% | 99.99% | 5 | 20% | 94% | 93% |
| 5 | 0.05% | 691 | 70.00% | 99.98% | 5 | 30% | 95% | 93% |
| 6 | 0.10% | 793 | 99.90% | 100.00% | 10 | 1% | 92% | 92% |
| 7 | 0.10% | 786 | 95.00% | 99.99% | 10 | 6% | 93% | 92% |
| 8 | 0.10% | 778 | 90.00% | 99.99% | 10 | 11% | 93% | 92% |
| 9 | 0.10% | 762 | 80.00% | 99.98% | 10 | 21% | 94% | 92% |
| 10 | 0.10% | 746 | 70.00% | 99.97% | 10 | 31% | 95% | 93% |
| 11 | 1.00% | 2117 | 99.90% | 100.00% | 100 | 7% | 79% | 79% |
| 12 | 1.00% | 2045 | 95.00% | 99.95% | 100 | 12% | 81% | 80% |
| 13 | 1.00% | 1970 | 90.00% | 99.90% | 100 | 17% | 82% | 80% |
| 14 | 1.00% | 1822 | 80.00% | 99.80% | 100 | 26% | 85% | 82% |
| 15 | 1.00% | 1673 | 70.00% | 99.70% | 100 | 35% | 88% | 83% |
| 16 | 3.00% | 4485 | 99.90% | 100.00% | 300 | 20% | 55% | 55% |
| 17 | 3.00% | 4296 | 95.00% | 99.85% | 300 | 24% | 59% | 57% |
| 18 | 3.00% | 4103 | 90.00% | 99.69% | 300 | 28% | 63% | 59% |
| 19 | 3.00% | 3717 | 80.00% | 99.38% | 300 | 36% | 70% | 63% |
| 20 | 3.00% | 3331 | 70.00% | 99.08% | 300 | 44% | 77% | 67% |

### Test performance and costs vs benefits of pooling

Tables 3 and 4 present test performance (negative and positive predictive values, false positive and false negative rates) for COVID-19 prevalence in the tested population of 0.05%-3%, and a range of test characteristics (sensitivity 70%-99.9%, specificity 70%-99.9%). As Table 3 demonstrates, changes in sensitivity do not have marked effects on negative predictive value. However, at higher prevalence, even small changes in the negative predictive value results in substantially more false negatives. Table 4 demonstrates that it is only with very high specificity (99.9%), and relatively high prevalence (>=1%) that most positive tests are true positives. In all other scenarios, most positives are false positives.

**Table 4:** Test performance and number of COVID-19 false positives per 10,000 population tested, for varied test specificities and disease prevalence (given a sensitivity of 99.9%)

| row | prevalence | Specificity | PPV | False positive rate | Number with disease in population/10,000 tested | Number of false positives per 10,000 tested |
| --- | --- | --- | --- | --- | --- | --- |
| 1 | 0.0005 | 0.999 | 33.32% | 0.10% | 5 | 10 |
| 4 | 0.0005 | 0.95 | 0.99% | 5.00% | 5 | 500 |
| 5 | 0.0005 | 0.9 | 0.50% | 10.00% | 5 | 1000 |
| 6 | 0.0005 | 0.8 | 0.25% | 20.00% | 5 | 2000 |
| 7 | 0.0005 | 0.7 | 0.17% | 30.00% | 5 | 3000 |
| 8 | 0.001 | 0.999 | 50.00% | 0.10% | 10 | 10 |
| 9 | 0.001 | 0.95 | 1.96% | 5.00% | 10 | 500 |
| 10 | 0.001 | 0.9 | 0.99% | 10.00% | 10 | 1000 |
| 11 | 0.001 | 0.8 | 0.50% | 20.00% | 10 | 2000 |
| 12 | 0.001 | 0.7 | 0.33% | 30.00% | 10 | 3000 |
| 13 | 0.01 | 0.999 | 90.98% | 0.10% | 100 | 10 |
| 14 | 0.01 | 0.95 | 16.79% | 5.00% | 100 | 500 |
| 15 | 0.01 | 0.9 | 9.17% | 10.00% | 100 | 1000 |
| 16 | 0.01 | 0.8 | 4.80% | 20.00% | 100 | 2000 |
| 17 | 0.01 | 0.7 | 3.25% | 30.00% | 100 | 3000 |
| 18 | 0.03 | 0.999 | 96.86% | 0.10% | 300 | 10 |
| 19 | 0.03 | 0.95 | 38.19% | 5.00% | 300 | 500 |
| 20 | 0.03 | 0.9 | 23.60% | 10.00% | 300 | 1000 |
| 21 | 0.03 | 0.8 | 13.38% | 20.00% | 300 | 2000 |
| 22 | 0.03 | 0.7 | 9.34% | 30.00% | 300 | 3000 |

### Mechanisms for enhanced surveillance

We highlight key public health system requirements and factors that may affect capacity to implement enhanced surveillance below:

#### Community engagement

The fever and cough surveillance strategy provides the opportunity for simple and clear messaging to the community: isolate and get tested if you have fever and cough. This must include all age groups and could be supported with a hotline and community-based support, and draw on the lessons learnt in regards to community engagement from this and current pandemic responses, both internationally and in Australia. (see Discussion for further details)

#### Sample collection and testing capacity

Where there is limited testing capacity, pooling of samples would reduce the number of tests required per individual to be tested (see Table 3). This is now being implemented in some settings.^15^ This will greatly reduce the need for testing reagents, especially in low prevalence settings. Promising strategies for further enhancing the coverage and feasibility of such testing include self-collection of samples, which has been found to have similar performance to health worker collected samples in initial research on COVID-19.^27^

If proven as reliable, this would reduce the number of health staff and PPE required for enhanced testing. Previous studies on influenza have found that self-collection of samples reduced sensitivity (87% sensitivity compared to clinician collected samples).^28^ As Table 3 demonstrates, even with such a reduction, overall benefits outweigh the costs in terms of false negatives if such reductions in performance are associated with increased testing capacity. Another promising strategy under review is testing of saliva samples, which will both allow self-collection, and also not require equipment such as swabs, which have been another limitation on testing capacity.

#### Epidemiological investigation and contact tracing

For enhanced surveillance to lead to pandemic control, all confirmed cases must be investigated. Contact tracing should be structured based on two groups of contacts: (a) upstream contacts, i.e. those who are the potential source of transmission to the identified case, and (b) downstream contacts, i.e. those likely to have had contact with the case while the case was infectious. Management will vary depending on which type of contact is identified:

a. Upstream contacts: intensive case finding and testing of these cases, whether symptomatic or not, is important, both with PCR-based testing to identify current infection, and with serology to identify past infection. The model indicates 2 or 3 cases (including the primary case) within the upstream transmission tree once it is detected at primary care with high surveillance (Figures 1 and 2). Full contact tracing of the entire transmission tree, which would include another 2-3 chains generated by these cases, would thus require around 22-33 contacts traced.29 It is important to note that these contacts could only be identified through upstream contact tracing.
b. Downstream contacts: optimal follow up includes intensive case finding and quarantining of these contacts, with home monitoring (twice daily temperature checks, GPS monitoring of compliance) and PCR testing for active disease once the 14-day quarantine and follow up period concludes to ensure they do not have asymptomatic disease. On average, 11 such contacts per case needing 14 days follow up per week would be expected.^29^

Although investigation of downstream contacts is part of most surveillance strategies, follow up of upstream contacts is not uniformly undertaken, but is a feature of those settings implementing successful control1,^30,31^ Investigation of contacts can be supported through training and integration of community volunteer networks into existing surveillance and contact tracing teams. Software may assist considerably in these efforts,^18^ and, linked to penalties for those found not to be with their phone during random spot checks by authorities, is again an important part of follow up in those settings with effective control.^32^

**Figure 2:**
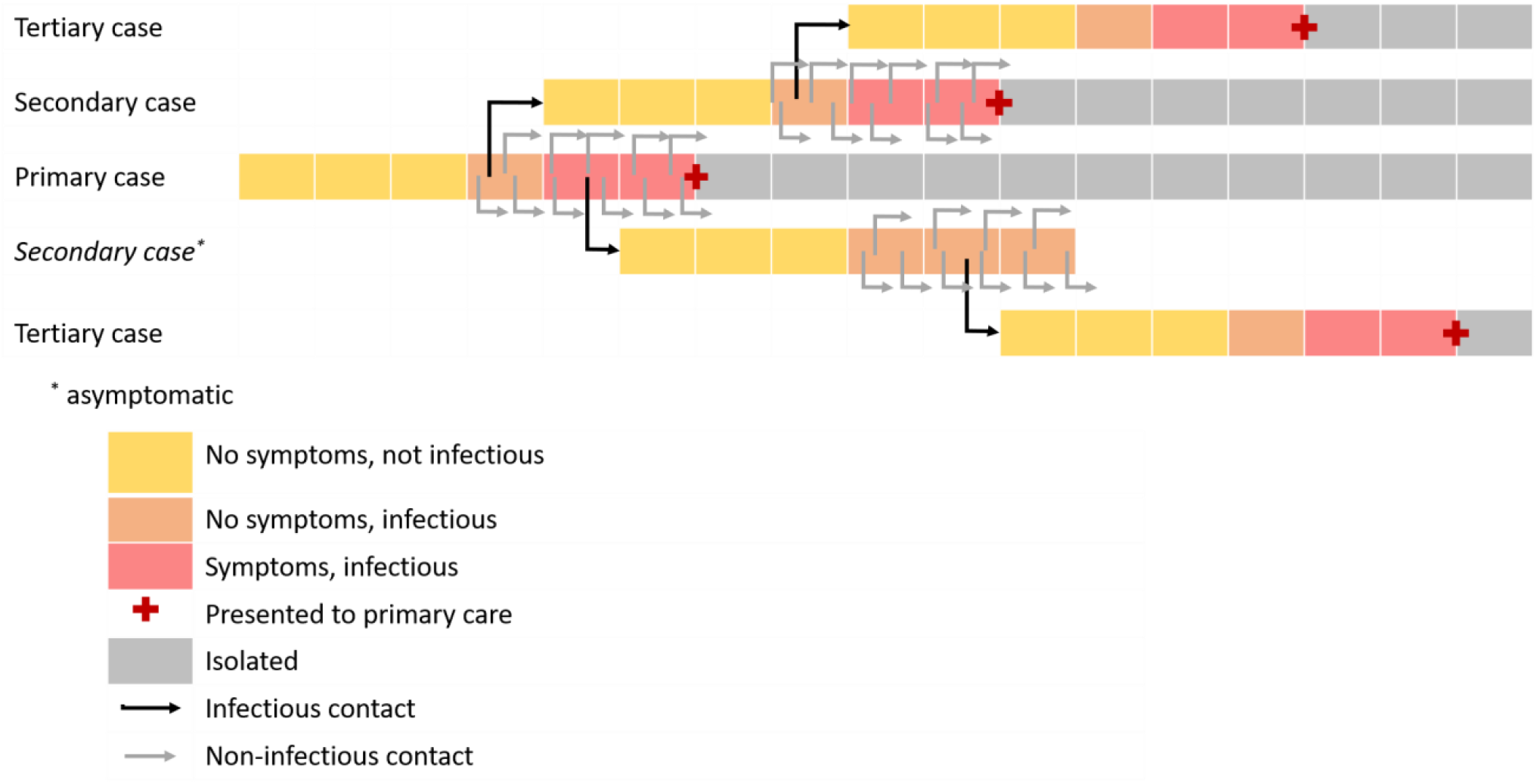
Upstream transmission tree at point of detection of case presenting to primary care.

#### Emerging strategies for enhanced surveillance

Recent studies have identified a range of symptoms associated with COVID-19. Symptoms such as anosmia and loss of taste appear to be relatively common in those with infection In a recent study based on ongoing community surveillance through online self-report of symptoms, the prevalence of these two symptoms was 59% in those who tested positive for COVID-19, and 19% in those who did not. ^33^ However, this study did not provide information on the prevalence of anosmia and loss of taste, without fever or cough, in the population testing negative for COVD-19 or in the general population. Once this data is available, it will be possible to assess the feasibility and added value of including anosmia and loss of taste in the surveillance case definitions.

Screening of wastewater for evidence of community transmission is another emerging strategy that is receiving increased attention^34^ and is another strategy that could provide a useful addition to primary care screening systems. It could also be used to monitor, on an ongoing basis, high risk facilities such as hospitals and aged care facilities in order to identify early transmission in such settings. It appears sensitive, and further assessment of such population-level environmental screening strategies is warranted. However, even if it proves sensitive, as it cannot identify specific infected individuals of chains of individuals, it must be linked to intensive human surveillance if it is to effectively support control of COVID-19 community transmission.

#### Indicators of success: Monitoring and evaluation of surveillance system performance

The following are recommended indicators for assessing the performance of enhanced COVID-19 surveillance:

System coverage, uptake and completeness:

- **100% fever and cough in the community screened for COVID-19:** weekly percent population screening target for all locations to be based initially on Table 1, varying with expected monthly total fever and cough incidence (i.e. 0.6%-3% of the population). This percent target should be reassessed monthly based on sentinel influenza surveillance systems (e.g. Flutracker, plus additional community-based surveys.) Surveillance system performance should also be validated through, for example, random community-based surveillance for unreported fever and cough at household and in general primary care settings. Fever and cough prevalence has decreased due to social distancing, however as these measures are lifted, it would be expected that prevalence will increase, especially if lifting occurring during winter.
- **The above indicator is the primary indicator to be monitored on an ongoing basis, regardless of whether phase of response is aiming towards elimination or maintaining elimination**.
- Related performance indicators:
  - 100% uptake of screening for fever and cough in sentinel surveillance populations, this can be part of the information collected during sentinel follow-up and monthly fever and cough prevalence surveys recommended above.
  - Community understanding of testing criteria, attitudes towards uptake of screening, practices related to screening, views on feasibility and burden, support services for enabling screening (again, can be included in sentinel surveillance and surveys)

Indicators of successful progress towards elimination of community transmission, given system coverage, uptake and completeness indicators have all been met:

- 100% of newly reported cases are travel related and/ or known contacts of confirmed cases.
- 100% of newly reported cases are tested on day of symptom onset.
- 100% of newly detected cases have been under quarantine from time of exposure event.
- 100% complete follow up of all contacts. Initially, as the modelling suggests, the number of upstream contacts under follow up per case should be at least 2 times number of downstream contacts under follow up (as there will be 2-3 upstream transmission branches for every identified case, and total number of contacts under follow up per case should be >35, unless there is a clear justification for lower. This figure should also be reviewed regularly, and contact case definitions updated based on serosurveys and screening for viral shedding around identified cases, including in high-risk settings (institutional settings, schools, health care facilities). Complete follow-up includes:
  - For upstream contacts: PCR and serological testing at time of case detection
  - For downstream contacts-documented quarantining for 14 days post last contact, linked to PCR testing at end of quarantine period to exclude asymptomatic viral shedding.
- 0% hospitalised new cases and/or deaths relative to total new community-acquired cases.^35^.

Indicators of successful elimination of community transmission, given system coverage, uptake and completeness indicators have all been met:

- 100% of new cases are travel related
- 0% of new cases are classified as unknown source or local community transmission-related exposure.

## Discussion

This study shows that timely detection and management of community transmission of COVID-19 is feasible. It shows that testing for infection in primary care patients presenting with cough and fever is an efficient, effective and feasible strategy for the detection and elimination of transmission chains.

### Application to surveillance scenarios

#### Increasing uptake of appropriate testing

Ensuring communities understand the need and value of presenting for screening is the fundamental requirement for successfully achieving and maintaining elimination of COVID-19 community transmission. This has been demonstrated both in countries which have high testing levels and have not seen resurgence despite lifting of containment measure, ^36^ and in those countries that successful controlled transmission through containment, but are seeing resurgence of community transmission due to limited testing.^37^ The fever and cough surveillance strategy provides the opportunity for simple and clear messaging to the community: isolate and get tested if you have fever and cough. In other settings, provision of paid sick leave for those with fever and cough supports compliance of testing and home isolation.^1^ Strategies for effective community engagement from other pandemics also provide guidance in this regard.^7,14^ Also of great relevance is the response of the Australian community to date, which has demonstrated a high uptake of highly burdensome containment measures.^4^ The community has also demonstrated a high uptake of testing each time testing criteria have been expanded.^38^ It is reasonable therefore to expect, that with a consistent and coordinated community engagement strategy and interventions that support timely and easy access to testing, uptake will be high.

#### Screening case definition: hospital vs symptoms vs asymptomatic

Reliance on hospital cases to signal transmission has lower sensitivity, with sensitivity decreasing as the reproductive number increases. It also has other disadvantages. Overall, the proportion of symptomatic cases hospitalised with COVID-19 is 20% globally,^21^ and the true proportion is likely to be lower if asymptomatic cases are included. A focus of increased hygiene, social distancing and containment measures has been on protecting those at most risk of severe disease and hospitalisation such as the elderly, immunocompromised or those with chronic respiratory or cardiovascular disease.^39,40^ If such measures successfully protect vulnerable populations, we would expect to see a substantial decrease in the proportion of cases with severe disease, and therefore the proportion hospitalised. This would further diminish the value of hospitalised disease as an indicator of transmission, as such cases may occur even later or not at all in many transmission chains.

Random testing of sentinel asymptomatic groups is inefficient at most levels of likely prevalence. The level of testing required to be confident of reliably detecting all transmission chains in the community, or obtaining precise estimates useful for policy making, is unlikely to be feasible on an ongoing basis in most settings. As Table 2 demonstrates, at a range of COVID-19 prevalence, and even with very high levels of testing, the majority of cases in the population are missed, and the estimates of prevalence have a comparatively wide range of uncertainly around them. It should also be noted that these results are for cross-sectional testing in the sentinel population, and would need to be repeated on an ongoing basis to constitute an effective surveillance system, entailing a high level of frequent, repeated and large-scale testing. Even if testing capacity were increased to the level at which the entire asymptomatic population could be screened, this could not be feasibly undertaken in a narrow time window. As a result, if there is undetected transmission, there is the potential for disease to move from those not tested to those previously tested negative over the time period in which the screening was underway.

#### Screening of contacts

Upstream contact tracing, including widespread testing of asymptomatic low and high risk contacts of cases, is the one situation where testing of asymptomatic cases is warranted. Currently, a proportion of all cases identified in Australia are classified as ‘unknown’ source of exposure.^5^ As Figure 2 demonstrates, this by definition means their source of exposure is likely to have initiated multiple chains of transmission by the time the downstream case is identified. Widespread testing of all contacts, upstream and downstream, including low risk contacts around such cases, and even geographical/fomite related testing of possible sources of infection is the most effective strategy for filling the gaps in transmission mapping indicated by such cases, and therefore ensuring identification and management of such unrecognised community transmission.

#### Limited testing capacity

Pooling is efficient, as it considerably increases the coverage of testing. Even if pooling results in a decrease in sensitivity of pooled tests, the overall number of cases in the population missed due to a higher number of false negatives is small compared to the number that would be missed due to not being tested when pooling is not applied. As Table 3 demonstrates, this is the case even if sensitivity following pooling were to drop to 70%, at COVID-19 prevalence ranging from 0.05%-3.If pooled testing is applied in low-prevalence settings with the expectation that sensitivity will be lower, messaging to the public must indicate that some cases may be missed, and that those with symptoms should isolate regardless of test results

#### Changes in specificity

As Table 4 demonstrates, at low prevalence, small changes in specificity result in large changes in the positive predictive value, and therefore most positive tests are false positives. At higher prevalence, the proportion of positive tests that are false positives decrease. Although specificity is considered high (100%) for PCR tests, the gold standard, the impact of reduced specificity is an important consideration when assessing rapid diagnostic tests coming to market. It may be that a contributor to the very high levels of asymptomatic disease reported in some settings may be partly due to lower test specificity, and this should be investigated further.

#### Expanded testing capacity, low prevalence

Ideally, under these conditions we would expand the case definition for screening. Inclusion of symptoms emerging as useful predictors of early and/or mild disease (e.g. anosmia and loss of taste)^33^ may increase the sensitivity of the system and allow earlier detection of transmission chains, especially once containment is lifted and the reproductive number for any new cases increases accordingly.

#### Impact of seasonality

The determining factor in regards to the number of individuals requiring testing is the prevalence of screening case definition symptoms in the general community. As Table 1 demonstrates, this means that at the peak of the influenza season, the testing requirements will increase between three to six fold compared to the non-influenza season. In Australia, peak influenza transmission occurs from late May-early July. In addition, if COVID-19 has a seasonal pattern, and this is likely to be the case considering other respiratory pathogens and similar SARS-CoV viruses,^41^ any residual transmission that continues into these months is likely to be amplified by seasonal effects. This suggests that given a strategy of strong containment and exhaustive surveillance aimed at elimination of COVID-19 community transmission, investing as much as possible in achieving this before the peak of the influenza season will optimise the value of every investment made in testing, contact tracing and containment.

It is important to note that our study only assessed surveillance and related measures for control of local transmission. One of the main contributors to transmission in Australia, and an important factor in effective control, has been how Australia’s borders, both air and sea, have been managed. Hotel quarantining of new international arrivals began on March 28^th^, ^42^ and appears to have contributed to the marked reduction in locally acquired disease, although disaggregation of data over time in relation to place of acquisition is not yet possible in the publically available COVID-19 data reported by the Australian Government. An enhanced surveillance system that has achieved elimination of detectable disease from the community is designed to detect and manage early re-introductions or emergence, including travel-related disease. However, continuation of effective border controls once other social distancing and control measures are lifted will reduce the pressure on the surveillance system to identify and manage imported disease and related local transmission once elimination has been achieved and while maintaining a relatively normal return to life within Australia.

The other major challenge to the surveillance system will come from superspreading events. Both during this outbreak and in past, significant outbreaks of other infectious diseases, such events centre around religious or burial activities,^43^ other large gatherings where contact occurs, and around health facilities and other institutional settings.^44^ Limiting the size and opportunities for transmission in such settings that are non-essential will be an important part of control. However, health facilities are essential services, and must continue functioning. Measures for heightened general infection prevention and control that cover both staff and patients, and deal not only with disinfection, but with patient and staff flow and which are linked to intensive surveillance, are a critical aspect of ongoing COVID-19 management in such settings even once elimination has been achieved. Such measures can and should also be applied to essential services and settings where there is a high proportion of vulnerable individuals (e.g. aged care), in the latter case not to prevent superspreading events, but to protect those most vulnerable to severe disease.

Australia currently has observed community transmission of COVID-19 in some jurisdictions, while in others, most cases are in or linked to returned travellers. Measures implemented at the end of March 2020 include isolation of cases, border controls, quarantine of contacts and social distancing. These measures have been escalated in some jurisdictions to include closure of schools, universities and non-essential workplaces.^29-34^ Testing rates are relatively high by international standards,^35^ and COVID-19 surveillance in some jurisdictions in Australia has moved beyond just testing of high-risk populations (e.g. symptomatic returned travellers and contacts of cases) to inclusion of testing of individuals likely to reflect community-based transmission (e.g. symptomatic health care workers and patients with pneumonia), and testing of fever and cough as proposed in this paper.^45^ This provides a foundation on which to bring about greater current pandemic control, as well as a means of detecting early resurgence in the community following successful control.

## Conclusion

Given our findings, we recommend exhaustive testing of patients presenting with fever and cough in primary care as the most efficient and feasible means of detecting all community transmission of COVID-19 in high and low transmission settings. This is in addition to current testing regimens such as the testing of symptomatic travellers, contacts, health care workers and hospitalised pneumonia cases. Once community cases are identified, detailed and meticulous upstream and downstream contact tracing, linked to quarantining of all contacts, and both antigen and serological testing of all upstream contacts who may be the source of infection, will support elimination of community transmission, and rapid control if and when reintroductions of disease occur. This strategy optimises the likelihood of remaining in the elimination phase while allowing for ongoing lifting of containment measures. Community engagement in order to ensure high levels of testing uptake and compliance with follow up measures in identified cases and their contacts is critical to successful implementation of this strategy.

## Data Availability

The data are readily available upon request.

## Contributors

KL and KG conceived of the original idea for the study. KL, KG, EB and TS searched and reviewed the available evidence. KL, KG and DO’D conducted the data analysis and modelling. All authors were involved in public health assessment of the surveillance and control options, interpretation, and writing of the report, and had final responsibility for the decision to submit for publication.

